# Contrastive Transformer-Driven Discovery of Temporal Hemodynamic Subphenotypes in Cardiac Surgery Patients

**DOI:** 10.64898/2026.03.27.26349519

**Authors:** Jacob M. Desman, Moein Sabounchi, Wonsuk Oh, Gagan Kumar, Ahmed Shaikh, Rohit Gupta, Umesh Gidwani, Anthony Manasia, Robin Varghese, John Oropello, Gordon Smith, Arash Kia, Prem Timsina, Ben Kaplan, Avniel Shetreat-Klein, Benjamin Glicksberg, Matthieu Legrand, Ashish K. Khanna, John A. Kellum, Patricia Kovatch, Roopa Kohli-Seth, Alexander W Charney, David Reich, Girish N. Nadkarni, Ankit Sakhuja

## Abstract

Cardiac surgery patients experience rapidly evolving hemodynamics in early post-operative period requiring intensive support. Identifying hemodynamic subphenotypes from these data can inform personalized management. Using 24-hour high-resolution physiologic and treatment data from 6,630 MIMIC-IV and 1,963 SICdb patients, we trained a transformer encoder with a reconstruction-contrastive objective to derive patient-level embeddings capturing multivariate temporal dynamics within first 24h of ICU stay and compared them against those generated by dynamic time warping (DTW). Spectral clustering uncovered three reproducible hemodynamic subphenotypes. Compared with subphenotype 1, subphenotype 3 received more IV fluids, vasopressors, inotropes, and exhibited higher in-hospital mortality (OR 5.85, 95 % CI 2.43-14.13), longer ICU stay (7.12 days, 95% CI: 5.52-8.73) and hospitalization (8.86 days, 95% CI: 6.57-11.16). DTW derived subphenotypes had weaker prognostic separation. Thus, contrastive-transformer framework identified more clinically meaningful temporal hemodynamic subphenotypes that may optimize post-operative risk stratification and inform personalized management.

## Introduction

With over 900,000 procedures performed annually, cardiac surgery is one of the most common surgeries in the United States^1^. Despite its routine nature, cardiac surgery carries significant risks. The early postoperative period, particularly in the first day after surgery, is highly dynamic, marked by rapid physiological changes requiring intensive monitoring and hemodynamic support. Managing this critical phase involves continuous assessment and titration of therapies to changing hemodynamics. The latter reflects the complex interplay between vital signs and therapeutic interventions that actively modulate them, including intravenous (IV) fluids, vasopressors, and inotropes. Maintaining optimal hemodynamics is crucial to minimize the risk of complications such as fluid overload and arrhythmias^2^^3^. Given the dynamic nature of the postoperative period, identifying hemodynamic subphenotypes that reflect distinct patterns of evolving relationships between vital signs and hemodynamic interventions could lead to valuable insights into patient trajectories and enable personalized management strategies.

Although patient heterogeneity has been documented in conditions such as sepsis and acute respiratory distress syndrome, similar studies among post-operative cardiac surgery patients remain limited^4–6^. Furthermore, existing approaches primarily rely on static data, overlooking crucial patterns embedded in high-frequency, time-series data collected during the early postoperative period^7–9^. Traditional clustering methods, such as k-means, are incompatible with time-series data and struggle with non-convex feature spaces^10,11^. Dynamic time warping (DTW) provides a distance metric that aligns sequences based on shape which allows for use in such traditional clustering algorithms, but fails to capture multivariate dependencies and precise temporal events across different features^12–14^.

Transformers, originally developed for natural language processing, offer a powerful framework for modeling long-range dependencies in sequential data through self-attention mechanisms^15^. Their ability to extract rich latent representations has made them increasingly relevant for analyzing complex, context-dependent physiological data in healthcare^16^. In parallel, contrastive learning is a self-supervised approach that enhances feature representations by explicitly differentiating between similar and dissimilar data points^17,18^. With its ability to encourage development of more structured and meaningful feature representations, it frequently outperforms other self- and semi-supervised learning approaches^17,18^. Combining transformers with contrastive learning leverages the strengths of both approaches where transformers can capture time-based interdependencies through self-attention while contrastive learning can enhance internal feature representations.

In this study, we apply transformers combined with contrastive learning to identify and validate hemodynamic subphenotypes in patients recovering from cardiac surgery. By modeling high-resolution, multivariate time-series data from the early postoperative period within the first 24 hours of ICU admission, our approach captures the temporal dynamics of physiological signals alongside the effects of therapeutic interventions. We hypothesize this will uncover clinically meaningful clusters reflecting distinct hemodynamic trajectories. Identifying these subphenotypes may offer valuable insights into patient hemodynamic profiles and support the development of more personalized postoperative management strategies.

## Results

### Population Characteristics

We identified 6,630 post-cardiac surgery patients from the Medical Information Mart for Intensive Care (MIMIC-IV) database to serve as the development cohort for model training and subphenotype identification. An external validation cohort of 1,963 post-cardiac surgery patients was drawn from the Salzburg Intensive Care Database (SICdb). Patients in the development cohort had a median age of 68 years (IQR: 61-76) and with 71.9% males (Table 1). Patients in the external validation cohort had a median age of 70 years (IQR: 60-75), with 73.3% males.

**Table 1.** Table of baseline population characteristics.

|  |  | MIMIC | SICDB | P-Value |
| --- | --- | --- | --- | --- |
| **n** |  | 6630 | 1963 |  |
| **Age (years), median [Q1,Q3]** |  | 68.0 [61.0,76.0] | 70.0 [60.0,75.0] | 0.162 |
| **Weight (kg), median [Q1,Q3]** |  | 83.4 [72.0,96.0] | 80.0 [70.0,90.0] | <0.001 |
| **Gender (Male), n (%)** | Male | 4764 (71.9) | 1438 (73.3) | 0.235 |
| **Race, n (%)** | Asian | 148 (2.2) |  |  |
|  | Black | 250 (3.8) |  |  |
|  | Hispanic | 191 (2.9) |  |  |
|  | Native American | 12 (0.2) |  |  |
|  | Other | 243 (3.7) |  |  |
|  | Unknown | 908 (13.7) |  |  |
|  | White | 4878 (73.6) |  |  |
| **NE Rate (mcg/kg/min), median [Q1,Q3]** |  | 0.008 [0.001,0.034] | 0.016 [0.004,0.058] | <0.001 |
| **Inotrope Rate (mcg/kg/min), median [Q1,Q3]** |  | 0.000 [0.000,0.000] | 0.000 [0.000,0.000] | <0.001 |
| **Heart Rate (bpm), median [Q1,Q3]** |  | 81.6 [77.0,87.4] | 80.0 [77.2,84.9] | <0.001 |
| **Systolic Blood Pressure (mmHg), median [Q1,Q3]** |  | 112.3 [107.8,117.2] | 107.6 [100.0,117.9] | <0.001 |
| **Diastolic Blood Pressure (mmHg), median [Q1,Q3]** |  | 56.4 [52.7,60.0] | 55.0 [51.2,59.4] | <0.001 |
| **Mean Arterial Pressure (mmHg), median [Q1,Q3]** |  | 72.7 [69.6,75.9] | 71.4 [67.8,77.5] | <0.001 |
| **Respiratory Rate (per min), median [Q1,Q3]** |  | 17.7 [16.2,19.3] | 14.9 [14.0,16.1] | <0.001 |
| **Temperature (Celsius), median [Q1,Q3]** |  | 36.7 [36.5,36.9] | 37.2 [36.9,37.4] | <0.001 |
| **SpO2 (%), median [Q1,Q3]** |  | 97.6 [96.7,98.5] | 98.1 [97.0,98.9] | <0.001 |
| **Total Fluid Intake (mL), median [Q1,Q3]** |  | 3994.4 [3124.3,5133.3] | 5403.2 [4338.4,7171.6] | <0.001 |
| **Total Fluid Output (mL), median [Q1,Q3]** |  | 2547.0 [1995.0,3214.7] | 2432.2 [1919.0,3060.8] | <0.001 |
| **Total Crystalloids (mL), median [Q1,Q3]** |  | 2942.4 [2283.7,3856.4] | 3914.8 [2810.7,5676.1] | <0.001 |
| **Total Colloids (mL), median [Q1,Q3]** |  | 500.0 [0.0,1000.0] | 0.0 [0.0,0.0] | <0.001 |

### Clustering Quality

The optimal number of subphenotypes (k) was determined to be 3 when clustering the contrastive transformer embeddings. This decision was informed by both the silhouette score and the Calinski-Harabasz index. Although the 2-subphenotype solution resulted in a marginally higher silhouette score (0.19 for 2 subphenotypes vs. 0.13 for 3 subphenotypes), indicating better within-cluster cohesion, it substantially oversimplified the inherent complexity as evidenced by over 95% of patients being assigned to a single subphenotype (Table 2)^19^. In contrast, the 3-subphenotype solution demonstrated superior between-cluster separation, as reflected by a substantially higher Calinski-Harabasz index (299.90 vs. 493.27 for 2 vs. 3 clusters). Consequently, we selected the 3-subphenotype solution as the more clinically meaningful and representative structure of the underlying data.

**Table 2.** Percentage of patients in the training set falling into cluster assignments. Silhouette score and Calinski-Harabasz index shown for each number of clusters.

|  | Contrastive | DTW | Contrastive | DTW | Contrastive | DTW | Contrastive | DTW | Contrastive | DTW | Contrastive | DTW |
| --- | --- | --- | --- | --- | --- | --- | --- | --- | --- | --- | --- | --- |
| Subphenotype Number |  |  |  |  |  |  |  |  |  |  |  |  |
| 1 | 95.39% | 94.89% | 64.04% | 94.59% | 64.90% | 94.46% | 34.78% | 94.46% | 32.32% | 93.28% | 34.20% | 76.64% |
| 2 | 4.61% | 5.11% | 31.37% | 2.76% | 27.64% | 2.71% | 29.43% | 2.22% | 25.27% | 3.28% | 23.08% | 17.04% |
| 3 |  |  | 4.59% | 2.65% | 4.59% | 2.80% | 28.36% | 1.92% | 20.92% | 1.96% | 16.96% | 2.82% |
| 4 |  |  |  |  | 2.87% | 0.02% | 4.61% | 1.38% | 14.01% | 1.42% | 10.41% | 2.00% |
| 5 |  |  |  |  |  |  | 2.82% | 0.02% | 4.61% | 0.04% | 9.05% | 1.42% |
| 6 |  |  |  |  |  |  |  |  | 2.87% | 0.02% | 3.47% | 0.04% |
| 7 |  |  |  |  |  |  |  |  |  |  | 2.84% | 0.02% |
| **Silhouette Score** | 0.19 | 0.87 | 0.13 | 0.84 | 0.13 | 0.83 | 0.13 | 0.82 | 0.12 | 0.75 | 0.11 | 0.22 |
| **Calinski-Harabasz Index** | 299.90 |  | 493.27 |  | 384.07 |  | 455.07 |  | 427.84 |  | 409.35 |  |

For comparison, we used k-means clustering with a DTW metric as a baseline (DTW k-means). DTW clustering also had a slightly higher silhouette score for the 2-subphenotype solution (0.87 for 2 subphenotypes vs. 0.84 for 3 subphenotypes), but in both cases over 90% of observations were consistently grouped into a single cluster - an imbalance that artificially inflated the silhouette scores, reflecting high within-cluster cohesion but limiting clinical meaningfulness. As the differences between the silhouette score and its impact on subphenotyping were minimal, we chose the 3-subphenotype DTW solution to maintain consistency and facilitate direct comparison with the contrastive transformer embedding-based subphenotypes (Table 2). Adopting a 3-subphenotype solution for both embedding methods also enabled consistent, clinically relevant, and methodologically rigorous comparisons.

Visual inspection of contrastive embedding spaces using principal component analysis (PCA) revealed a reproducible low-dimensional structure while t-distributed stochastic neighbor embedding (t-SNE) showed coherent local groupings, supporting the internal consistency and plausibility of the identified subphenotypes (Fig. 1). As DTW k-means operates on the raw time series and lacks a learned embedding space, these evaluations were only performed for contrastive embedding-derived subphenotypes.

**Figure 1.**
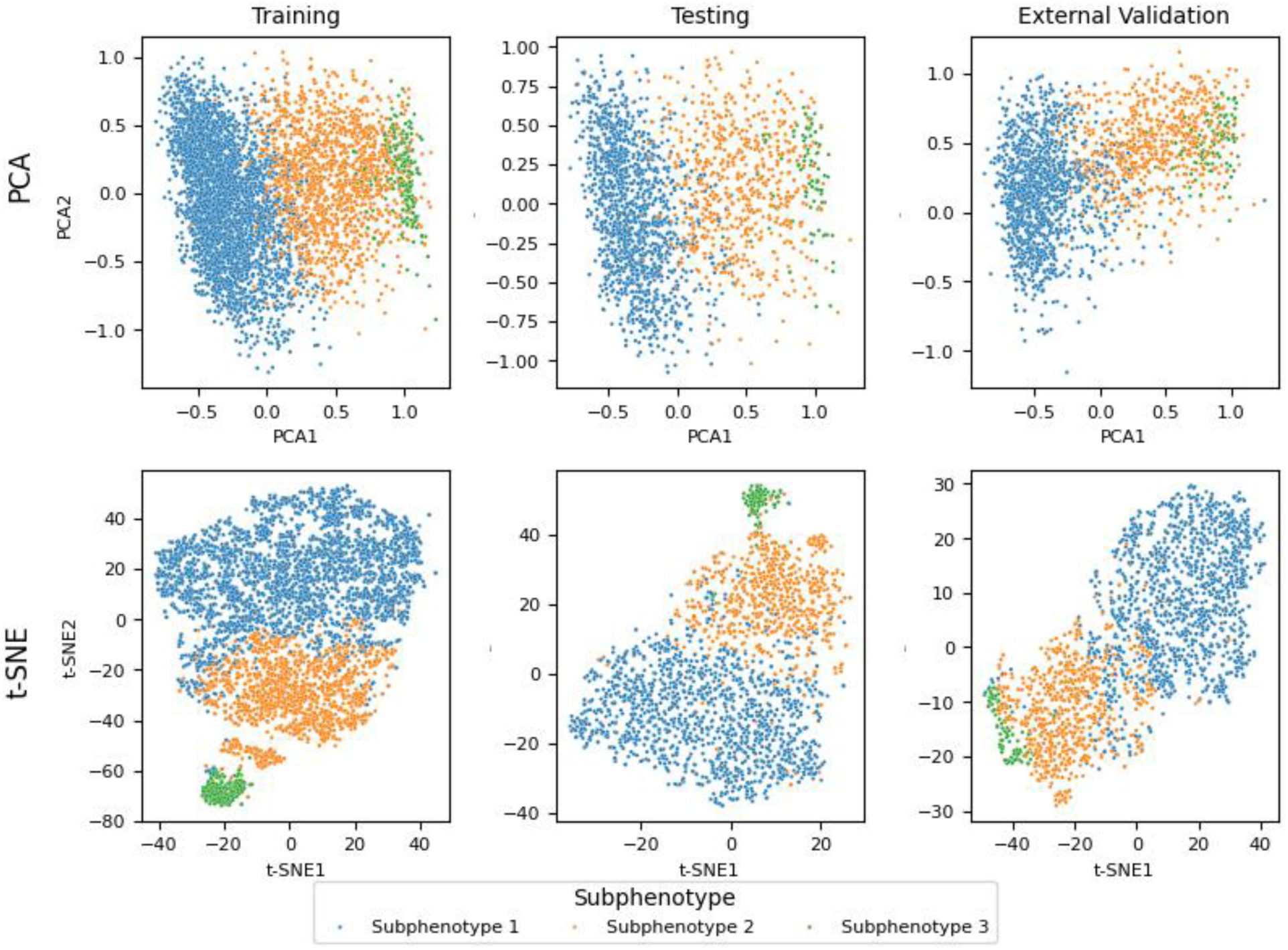
Low dimensional embedding representations. PCA and t-SNE embeddings of the transformer derived embeddings.

### Subphenotype Characteristics

There were no clinically meaningful differences in age or gender among subphenotypes across the datasets. There was a consistent, progressive increase in 24-hour total fluid intake, crystalloid administration, colloid administration, vasopressor doses, and inotropic doses from subphenotype 1 (low-acuity) to subphenotype 3 (high-acuity) (Table 3). Similar trends emerged within the DTW-derived clusters, however, these clusters contained markedly smaller intermediate and high acuity groups suggesting potential limitations in their clinical interpretability and practical utility (Supplementary Table 1, Supplementary Table 2).

**Table 3.**
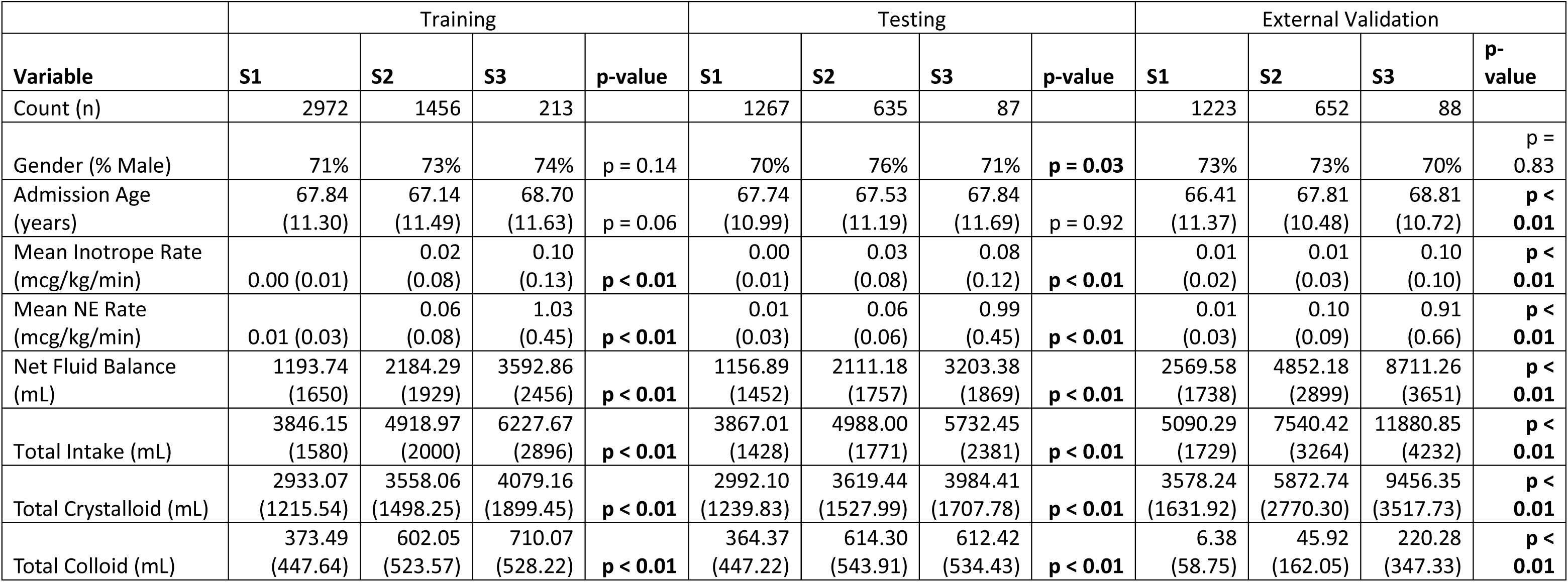
Clinical characteristics of patients assigned to each subphenotype. Subphenotypes derived from the transformer-derived embeddings. NE = norepinephrine equivalent, S1 = subphenotype 1, S2 = subphenotype 2, S3 = subphenotype 3

Analysis of hourly trends revealed distinct physiological trajectories across subphenotypes, with significant differences observed in the training, testing, and external validation datasets (Fig. 2). Subphenotype 1 represented a comparatively hemodynamically stable profile characterized by higher mean arterial pressures (MAP) and systolic blood pressures (SBP), lower total fluid intake, and lower utilization of crystalloids, colloids, vasopressors, inotropic support. In contrast, subphenotype 3 represented a high-acuity profile, with significantly lower MAPs and higher IV fluid administration, vasopressor and inotropic support (all p < 0.05; Fig. 2). Subphenotype 2 demonstrated an intermediate profile, with MAPs lower than subphenotype 1 but higher than subphenotype 3, and moderate but statistically significant utilization of IV fluids, vasopressors and inotropes (p < 0.05, Fig. 2). Parallel analysis using the DTW-derived subphenotypes revealed similar overall trends, however had less consistent separation and higher variance (Supplementary Table 1, Supplementary Fig. 2). Moreover, DTW-based clustering identified fewer statistically significant time-varying differences, particularly among vital signs, in both the held-out testing and external validation sets. This suggests that DTW-based clustering may overemphasize therapeutic interventions at the expense of capturing meaningful physiological distinctions, thereby limiting its overall clinical utility (Supplementary Fig. 2).

**Figure 2.**
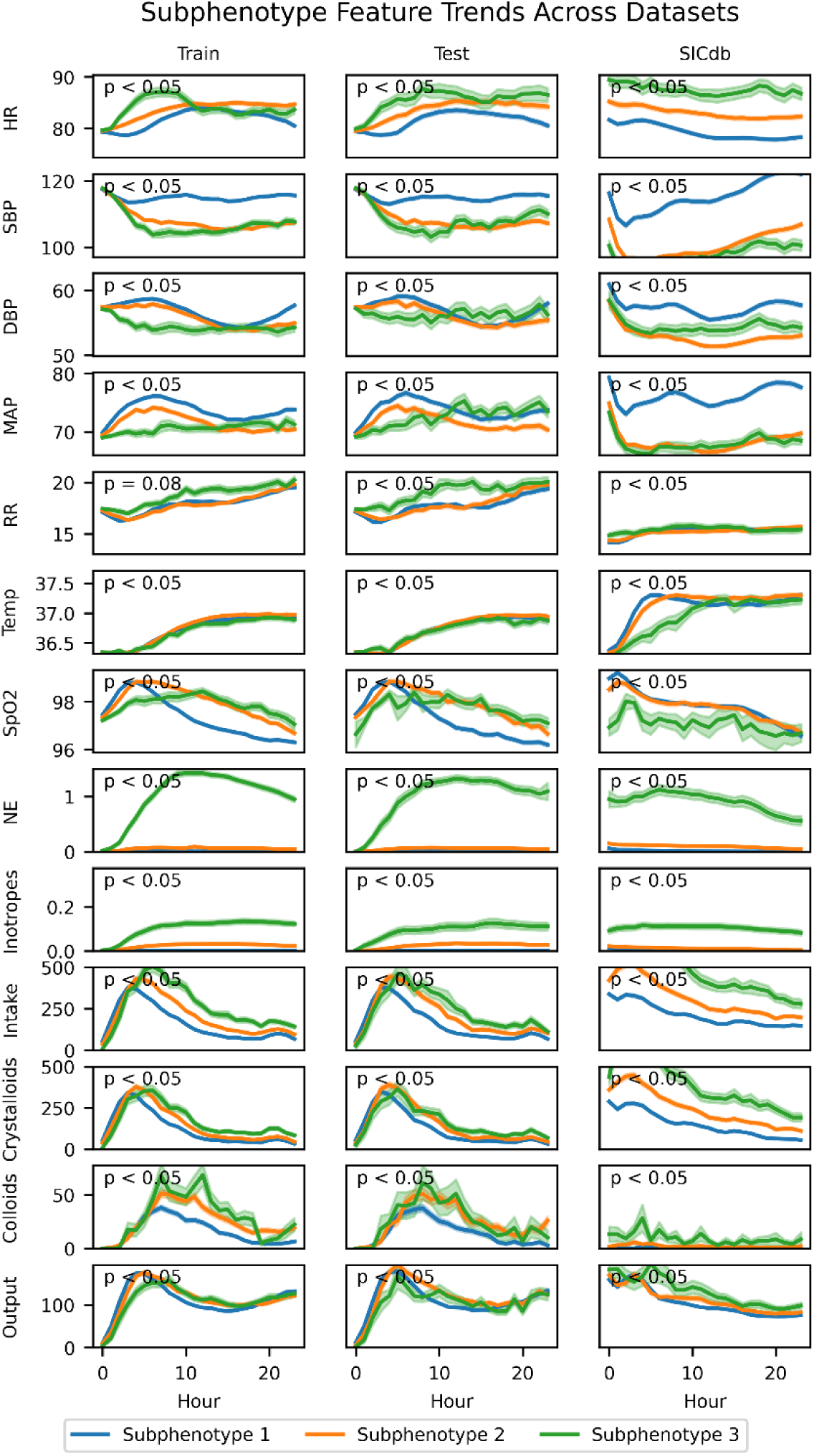
Features over time. Mean feature value ± standard error of the mean at each hour within each transformer-derived cluster. Linear mixed-effects modeling was used to test whether the time-series trend for each clinical feature differed significantly across patient subphenotypes. NE = norepinephrine equivalent dose; Inotropes = milrinone equivalent dose.

### Subphenotype Outcomes

We first evaluated clinical outcomes across subphenotypes using univariable analyses, which demonstrated a clear stepwise increase in mortality, ICU length of stay (LOS), and hospital LOS from subphenotype 1 to subphenotype 3 (all p < 0.01) (Table 4, Supplementary Table 2). To further assess the prognostic relevance of these subphenotypes, we performed adjusted analyses using logistic regression for mortality and linear regression for length of stays (Fig. 3). Compared to subphenotype 1, subphenotype 3 demonstrated the highest odds of mortality (training: OR 9.32, 95% CI: 3.86–23.82, p < 0.01; testing: OR 8.61, 95% CI: 1.10–72.99, p < 0.05; external validation: OR 5.85, 95% CI: 2.43–14.13, p < 0.01). Subphenotype 2 also showed a significantly elevated mortality risk relative to subphenotype 1, though lower than subphenotype 3 (training: OR 3.21, 95% CI: 1.52–7.16, p < 0.01; testing: OR 6.45, 95% CI: 1.71–34.88, p < 0.01; external validation: OR 3.08, 95% CI: 1.60–6.24, p < 0.01). Similar trends were observed for hospital and ICU LOS relative to subphenotype 1, with subphenotype 3 having the longest hospital LOS (training: 7.05 days, 95% CI: 6.18–7.91, p < 0.01; testing: 7.11 days, 95% CI: 5.80–8.43, p < 0.01; external validation: 8.86 days, 95% CI: 6.57–11.16, p < 0.01) and ICU LOS (training: 4.66 days, 95% CI: 4.10–5.22, p < 0.01; testing: 4.86 days, 95% CI: 3.97–5.76, p < 0.01; external validation: 7.12 days, 95% CI: 5.52–8.73, p < 0.01). Subphenotype 2 had intermediate LOS outcomes, consistently longer than subphenotype 1 but shorter than subphenotype 3 (hospital LOS: training 1.28 days, testing 1.91 days, external validation 2.89 days; ICU LOS: training 1.08 days, testing 1.39 days, external validation 2.25 days; all p < 0.01).

**Figure 3.**
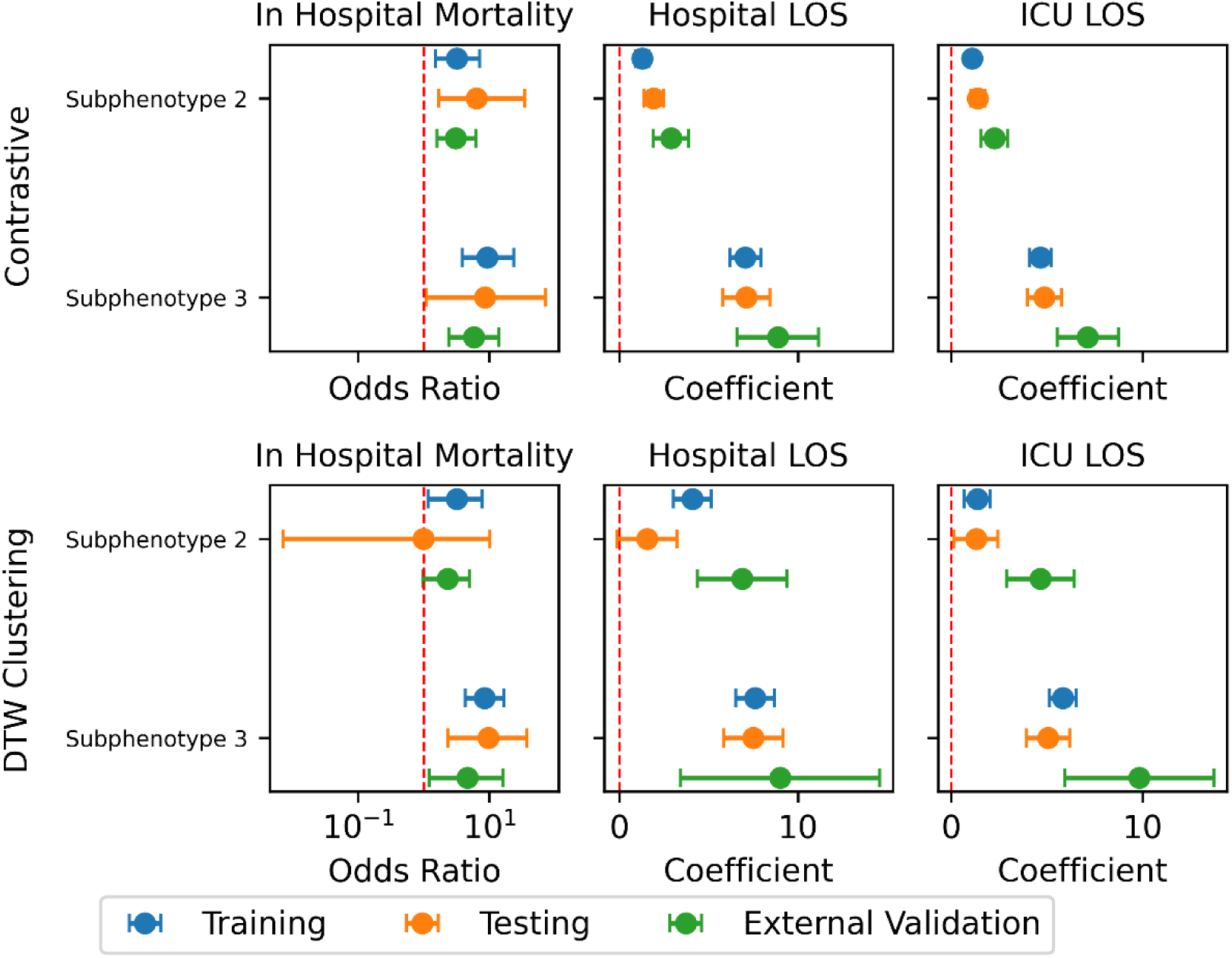
Outcome ORs. Adjusted OR for hospital mortality, and adjusted regression coefficients comparing hospital and ICU length of stay among patient subphenotypes across cohorts. 95% CI shown. Analyses adjusted for baseline demographics (age, gender) and organ dysfunction (maximum 24-hour SOFA score).

**Table 4.** Univariable analysis of outcome characteristics of patients assigned to each subphenotype. Subphenotypes derived from the transformer-derived embeddings. S1 = subphenotype 1, S2 = subphenotype 2, S3 = subphenotype 3.

|  | Training |  |  |  | Testing |  |  |  | External Validation |  |  |  |
| --- | --- | --- | --- | --- | --- | --- | --- | --- | --- | --- | --- | --- |
| Variable | S1 | S2 | S3 | p-value | S1 | S2 | S3 | p-value | S1 | S2 | S3 | p-value |
| Count (n) | 2972 | 1456 | 213 |  | 1267 | 635 | 87 |  | 1223 | 652 | 88 |  |
| ICU LOS (days) | 2.20<br>(2.77) | 3.55<br>(4.29) | 8.42 (9.82) | **p < 0.01** | 2.08<br>(2.13) | 3.77<br>(5.41) | 8.79<br>(9.23) | **p < 0.01** | 4.12<br>(4.92) | 7.58<br>(9.32) | 14.76<br>(13.20) | **p < 0.01** |
| Hospital LOS (Days) | 7.99<br>(4.87) | 9.71<br>(6.25) | 17.48<br>(13.15) | **p < 0.01** | 7.73<br>(4.19) | 10.11<br>(7.54) | 17.64<br>(11.17) | **p < 0.01** | 11.35<br>(7.80) | 15.63<br>(12.46) | 24.18<br>(18.16) | **p < 0.01** |
| In-Hospital Mortality (% Died) | 0% | 2% | 13% | **p < 0.01** | 0% | 1% | 6% | **p < 0.01** | 1% | 5% | 20% | **p < 0.01** |

Comparable but less consistent trends were observed for the DTW-derived subphenotypes. While mortality, hospital LOS, and ICU LOS generally increased with subphenotype acuity, confidence intervals were wider and significance was not uniformly observed across datasets (Fig. 3). These findings suggest that DTW-based subphenotypes may have lower clinical discriminability and prognostic relevance than those identified through contrastive learning (Fig. 3).

## Discussion

In this study, we applied a contrastive transformer framework to high-resolution, multivariate time-series data collected during the first 24 hours after cardiac surgery and identified three hemodynamic subphenotypes with clear differences in treatment intensity and outcomes. Unlike traditional clustering approaches that rely on static snapshots or shape-based distances, our model learns rich latent representations that capture both temporal dynamics and interdependencies among vital signs and interventions. The result is subphenotypes that are reproducible across internal and external cohorts and meaningfully risk stratify patients.

Our transformer-based approach for subphenotyping leverages transformer to encode long-range dependencies in multivariate time series^12^. It also employs contrastive learning to differentiate patient trajectories^14^. In contrast, DTW-based k-means clustering aligns sequences solely based on shape, missing complex interdependencies^17^. Consequently, DTW-based clustering assigned over 90% of patients to a single cluster, artificially inflating silhouette scores and obscuring meaningful distinctions. Additionally, DTW-based clusters showed less consistent separation in both held-out testing and external validation cohorts. These clusters had wider confidence intervals and fewer significant differences in vital signs and interventions over time. In contrast, subphenotypes derived from contrastive transformer embeddings demonstrated clear, reproducible separation of hemodynamic profiles and stronger prognostic relevance, with more precise risk stratification for mortality and length of stay across datasets. This highlights that latent representations learned via self-attention and contrastive objectives effectively capture clinically meaningful multivariate dynamics.

Our findings extend prior efforts in critical care subphenotyping by demonstrating that contrastive learning can reveal clinically plausible trajectories in the postoperative setting^20–23^. We also build upon previous work employing contrastive learning with transformer architectures that depend on well-defined index events, which may overlook important continuous physiologic patterns^24^. In contrast, our approach introduces an event-free, dual contrastive-reconstruction transformer that leverages the full postoperative hemodynamic data stream without requiring nuanced preprocessing. This design enables the model to capture subtle, real-time interactions between vital signs and therapeutic interventions that might otherwise remain under characterized. We found that whereas subphenotype 1 showed stable blood pressures and minimal fluid or vasoactive support; subphenotype 3 required aggressive interventions and experienced the highest mortality and length of stay; subphenotype 2 lay in between. These patterns held across MIMIC-IV and SICdb, suggesting that our approach generalizes beyond a single center. Moreover, our unsupervised method discovered clusters well aligned with prior clinical hypothesis driven studies on vasoactive and inotropic medication use, and postoperative fluids, further supporting the relevance of these subphenotypes^2,25–28^.

These temporal hemodynamic clusters enable risk stratification to identify high-acuity patients that are at risk for worse outcomes and need more intensive monitoring and resource utilization. On the other hand, identifying low-acuity patients will support safer de-escalation and earlier mobilization. Assigning patients to high, intermediate, or low-acuity groups allows proactive ICU bed planning, staffing allocation, and equipment scheduling. Finally, these well-defined subphenotypes create enriched cohorts for clinical trials, improving recruitment efficiency and statistical power for studies of postoperative management and hemodynamic therapies.

There are limitations that should be considered when interpreting this study. As a retrospective analysis, our findings are susceptible to unmeasured confounding. However, we validated our subphenotypes in an independent international cohort and adjusted outcome analyses for key confounders, supporting generalizability of our results. Second, these subphenotypes are based exclusively on the hemodynamic data from the first 24 hours of the post-operative period. This design enables rapid stratification but may miss clinically important trajectory changes that occur later in the ICU stay. Finally, we did not have data regarding intra-operative hemodynamic monitoring but its incorporation in future studies could further refine these subphenotypes.

In conclusion, this contrastive transformer framework discovered three distinct, clinically meaningful hemodynamic subphenotypes among patients after cardiac surgery. Identifying these subphenotypes paves the way for individualized hemodynamic management in this high-risk population. Furthermore, the contrastive transformer methodology for subphenotyping offers a scalable, data-driven approach for uncovering meaningful time-series subphenotypes across other diseases, thereby advancing precision medicine and streamlining the design of targeted interventional trials.

## Methods

### Study Design and Databases

This was a retrospective study where we used the Medical Information Mart for Intensive Care (MIMIC-IV) database as the development cohort for model training and identification of subphenotypes^29^. MIMIC-IV is a single-center database comprised of over 200,000 deidentified ICU admissions from the Beth Israel Deaconess Medical Center from 2008-2019. We utilized the Salzburg Intensive Care Database (SICdb) as the international external validation cohort. This dataset provides highly granular physiological data from over 27,000 ICU admissions to the University Hospital Salzburg^30^.

### Study Population

We included adult patients (age ≥ 18) who were admitted to the ICU after cardiac surgery. In MIMIC-IV, we identified patients using the relevant International Classification of Diseases-10-Procedure Coding System (PCS) codes for cardiac surgery (Supplementary Table 3). As SICdb does not contain PCS codes, we identified cardiac surgery patients in this cohort based on their first ICU admission following surgery, as indicated by the “heartsurgerybeginoffset” variable in the dataset. We excluded patients with fewer than 24 hours of ICU data, those without a recorded weight, and those who died within 24 hours. A CONSORT diagram describing patient selection process is available in Supplementary Figure 1.

### Feature Extraction and Preprocessing

We extracted relevant patient demographics (age and sex) and routinely collected vital signs (heart rate, blood pressures, respiratory rate, temperature, and oxygen saturation) within first 24 hours after ICU admission. We also extracted data regarding intravenous (IV) fluids administered (both crystalloids and colloids), total input (including IV fluids, blood products, any oral intake, fluids from medications, etc.), total output (including urine output, ultrafiltration, chest tube output, drain outputs, vomiting, etc.), net fluid balance (as a difference between total input and total output), vasopressor amounts (dopamine, epinephrine, norepinephrine, phenylephrine, and vasopressin), and inotrope amounts (dobutamine and milrinone) within first 24 hours of ICU admission. We converted the vasopressor doses into norepinephrine equivalent dose^31^. Similarly, we converted the inotrope doses into milrinone equivalent dose^32^.

Data were aggregated into 1-hour windows. If multiple measurements were available for a feature within a single hour, we used either the mean (e.g. for vital signs) or the sum (e.g. for medication doses and fluid amounts), as appropriate^33^. Forward-fill imputation was employed for hours with missing data to carry forward the last known value, reflecting the most up to date knowledge available to clinicians^33^. Remaining missing values were imputed with k-nearest neighbors (k=5).

### Model Architecture

We implemented the model’s encoder using a transformer architecture which employs a learnable self-attention mechanism that allows every feature and timestep, regardless of temporal distance, to contribute to the internal representations based on relevance^15^. Our embedding strategy is inspired by SimCLR, a contrastive learning framework that maps data into a latent embedding space using a projection head composed of a single hidden layer multi-layer perceptron (MLP)^18^. As Chen et al (2020) observed that the layer immediately preceding the projection head retains more informative representations, as the projection head may induce information loss related to compression, we applied mean pooling across timesteps at the preceding layer to generate a single latent vector per patient^18,34^. This approach preserves semantically rich features while summarizing the patient’s temporal data into a unified embedding^34^. Additionally, we introduced a reconstruction branch to ensure the embeddings comprehensively encode patient states. All model hyperparameters can be found in Supplementary Table 4.

### Model Training

The MIMIC-derived development cohort was divided into training (70%) and held out testing (30%) sets. We trained the model on the training set for 10,000 epochs until loss was no longer observed to decrease before using the Adam optimizer with an initial learning rate of 3.0 × 10⁻^4^ with a cosine annealing with warm restarts learning rate scheduler. Input features were normalized using robust z-score normalization.

A dual objective loss as follows:

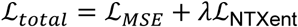

with ℒ_*MSE*_ defined as:

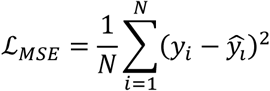

and normalized temperature-scaled (NT-Xent) contrastive loss defined as:

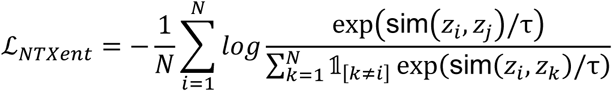

with sim(⋅,⋅) denoting cosine similarity and τ the temperature parameter^17^.

This dual objective encourages the model to reconstruct the original time series (mean square error (MSE) loss) while learning discriminative embeddings (NT-Xent loss)^17,18^. Colloquially, patients with similar clinical trajectories are pulled closer in latent space while those with dissimilar trajectories are pushed apart. All models were trained using Python 3.10.4 and PyTorch^35^.

While our contrastive learning strategy is inspired by the framework proposed by Chen et al, which maximizes agreement between augmented views of the same input, there are key distinctions in our implementation^18^. First, Chen et al.’s work was developed for image data using convolutional neural networks, whereas we apply contrastive learning to longitudinal clinical time series using a transformer-based encoder. Second, unlike the original framework, we incorporate a dual-objective loss that combines a reconstruction objective (MSE loss) with the contrastive NT-Xent loss. This allows our model to not only learn discriminative embeddings but also retain clinically relevant features necessary to reconstruct all aspects of the input sequence. Third, we conduct contrastive learning on time series rather than imaging data. To our knowledge, this is the first application of contrastive learning to clinical time series using the two augmented views approach with a transformer architecture and reconstruction-contrastive objective.

We restricted the augmentations to small, clinically plausible modifications to avoid distorting critical content within the time series. Specifically, we randomly applied up to 4 composable transformations which were jitter, which adds Gaussian noise drawn from N(0, 0.2^2^) to each time step; scaling, which multiplies each time step by a random factor drawn from N(1, 0.1^2^); temporal shifting, which shifts the entire sequence forward or backward by a few steps and fills vacant positions with zeros; and time masking, which randomly zeroes out a step of the sequence. These augmentations were chosen to embed salient clinical features while remaining robust to noise and irregularities in data recording.

### Subphenotype Development

We next applied unsupervised machine learning to the learned embeddings to identify naturally emerging clusters. Spectral clustering was chosen for its effectiveness in detecting non-convex, irregularly shaped clusters of varying sizes^36^. Therefore, spectral clustering was applied to the training set, and nearest neighbors was used to categorize embeddings from internal validation and external validation into those groups.

Determining the optimal number of clusters *k* remains a complex problem for which no universally accepted solution exists. We therefore calculated multiple metrics to inform this decision, including the silhouette coefficient and the Calinski-Harabasz index, across multiple values of *k*, and chose the number of clusters which balances metric performance while retaining clinical interpretability^37,38^. Alternative heuristic approaches, such as the commonly employed “elbow method”, rely heavily on subjective interpretation, motivating our use of more objective metrics before assessing for clinical interpretability. Therefore, we used silhouette coefficients and Calinski-Harabasz index to inform the number of clusters.

### Statistical and Analysis Plan

We compared baseline population characteristics using Kruskal-Wallis tests for continuous variables and Chi-squared tests for categorical variables. Characteristics of patient subphenotypes were compared using two-sided t-tests and Chi-squared tests as appropriate. To qualitatively assess the geometry of the learned embedding space and verify that the contrastive model produced meaningful subphenotypes, we generated two complementary two-dimensional visualizations. First, we applied principal component analysis (PCA) to capture the dominant axes of global variance and check for a reproducible low-dimensional structure^39^.

Second, we used t-distributed stochastic neighbor embedding (t-SNE) to preserve local neighborhood relationships and highlight fine-grained groupings^40^. Together, these plots enabled a rapid visual screen for coherent clusters: PCA demonstrated a stable overarching configuration, whereas t-SNE revealed tightly knit local clusters that corresponded to the three data-driven subphenotypes identified downstream.

We then used mixed-effects models to evaluate trends of vital signs, fluid balance, vasopressor doses, and inotrope dosing across the subphenotypes. Our primary outcomes included ICU length of stay, hospital length of stay, and in-hospital mortality. To account for potential confounding by baseline demographics and illness severity, we performed linear regression and Firth’s logistic regression analyses adjusting for age, gender, and maximum 24-hour SOFA scores^41^. We used Firth’s logistic regression as standard maximum likelihood logistic regression can produce biased estimates and unreliable confidence intervals when analyzing rare events as in our study^41,42^.

## Acknowledgements

This study was supported by NIH/NIDDK grant 5K08DK131286 (AS). This work was supported in part through the computational and data resources and staff expertise provided by Scientific Computing and Data at the Icahn School of Medicine at Mount Sinai and supported by the Clinical and Translational Science Awards (CTSA) grant UL1TR004419 from the National Center for Advancing Translational Sciences. Research reported in this publication was also supported by the Office of Research Infrastructure of the National Institutes of Health under award number S10OD026880 and S10OD030463. The content is solely the responsibility of the authors and does not necessarily represent the official views of the National Institutes of Health.

## Author Contributions

**CRediT taxonomy author statement:** Conceptualization – JMD, AS; Data curation – JMD, MS, AS; Formal analysis – JMD, MS; Funding acquisition – GNN, AS; Methodology – JMD, AS; Resources – GNN, AS; Software – JMD, AS; Supervision – GNN, AS; Writing – Original Draft Preparation – JMD, AS; Writing – Review & Editing – All authors contributed to review and editing. All authors contributed to the critical revision of the manuscript for important intellectual context. All authors have read and approved the manuscript.

## Code Availability

Code will be made available upon reasonable request to the corresponding author.

## Data Availability

All datasets used and analyzed in this present study are publicly available. Each dataset can be found in its associated online repository (MIMIC-IV: https://physionet.org/content/mimiciv/2.2/; SICdb: https://physionet.org/content/sicdb/1.0.8/).

## Notes

### Competing Interest Statement

A.S. is a consultant for Roche Diagnostics Corporation. G.N.N. is a founder of Renalytix, Pensieve, Verici, provides consultancy services to AstraZeneca, Reata, Renalytix, Siemens Healthineer and Variant Bio, and serves as a scientific advisory board member for Renalytix and Pensieve. He also has equity in Renalytix, Pensieve and Verici. J.A.K. reports receiving consulting fees from Astute Medical/bioMerieux, Astellas, Alexion, Chugai Pharma, Novartis, Mitsubishi Tenabe and GE Healthcare and is a full time employee of Spectral Medical. All remaining authors have declared no conflicts of interest.

### Author Declarations

All datasets used and analyzed in this present study are publicly available. Each dataset can be found in its associated online repository (MIMIC-IV: https://physionet.org/content/mimiciv/2.2/; SICdb: https://physionet.org/content/sicdb/1.0.8/). The SICdb data is deidentified.

## References

1. Virani, S.S., et al. Heart Disease and Stroke Statistics-2020 Update: A Report From the American Heart Association. Circulation 141, e139–e596 (2020).

2. Nielsen, D.V., et al. Health outcomes with and without use of inotropic therapy in cardiac surgery: results of a propensity score-matched analysis. Anesthesiology 120, 1098–1108 (2014).

3. Koponen, T., et al. Vasoactive-inotropic score and the prediction of morbidity and mortality after cardiac surgery. Br J Anaesth 122, 428–436 (2019).

4. Milam, A.J., et al. Derivation and validation of clinical phenotypes of the cardiopulmonary bypass–induced inflammatory response. Anesthesia & Analgesia 136, 507–517 (2023).

5. Komorowski, M., Green, A., Tatham, K.C., Seymour, C. & Antcliffe, D. Sepsis biomarkers and diagnostic tools with a focus on machine learning. EBioMedicine 86(2022).

6. Tran, T.K., et al. A systematic review of machine learning models for management, prediction and classification of ARDS. Respiratory Research 25, 232 (2024).

7. Milam, A.J., et al. Derivation and Validation of Clinical Phenotypes of the Cardiopulmonary Bypass-Induced Inflammatory Response. Anesth Analg 136, 507–517 (2023).

8. Avanzolini, G., Barbini, P. & Gnudi, G. Unsupervised learning and discriminant analysis applied to identification of high risk postoperative cardiac patients. Int J Biomed Comput 25, 207–221 (1990).

9. Zhao, X., et al. Machine learning approach identified clusters for patients with low cardiac output syndrome and outcomes after cardiac surgery. Front Cardiovasc Med 9, 962992 (2022).

10. Baek, M.S., Kim, J.H. & Kwon, Y.S. Cluster analysis integrating age and body temperature for mortality in patients with sepsis: a multicenter retrospective study. Sci Rep 12, 1090 (2022).

11. Hu, C., Li, Y., Wang, F. & Peng, Z. Application of Machine Learning for Clinical Subphenotype Identification in Sepsis. Infectious Diseases and Therapy 11, 1949–1964 (2022).

12. Bhavani, S.V., et al. Comparison of time series clustering methods for identifying novel subphenotypes of patients with infection. J Am Med Inform Assoc 30, 1158–1166 (2023).

13. Xu, Z., et al. Sepsis subphenotyping based on organ dysfunction trajectory. Crit Care 26, 197 (2022).

14. Su, C., et al. Identifying organ dysfunction trajectory-based subphenotypes in critically ill patients with COVID-19. Sci Rep 11, 15872 (2021).

15. Vaswani, A., et al. Attention is all you need. Advances in neural information processing systems 30(2017).

16. Zhou, H.Y., et al. A transformer-based representation-learning model with unified processing of multimodal input for clinical diagnostics. Nat Biomed Eng 7, 743–755 (2023).

17. Sohn, K. Improved deep metric learning with multi-class n-pair loss objective. Advances in neural information processing systems 29(2016).

18. Chen, T., Kornblith, S., Norouzi, M. & Hinton, G. A simple framework for contrastive learning of visual representations. in International conference on machine learning 1597-1607 (PmLR, 2020).

19. Xu, S., et al. Reviews on determining the number of clusters. Applied Mathematics & Information Sciences 10, 1493–1512 (2016).

20. Johnson, A.E.W., et al. Machine Learning and Decision Support in Critical Care. Proceedings of the IEEE 104, 444–466 (2016).

21. Vranas, K.C., et al. Identifying Distinct Subgroups of ICU Patients: A Machine Learning Approach. Crit Care Med 45, 1607–1615 (2017).

22. Zweck, E., et al. Phenotyping Cardiogenic Shock. J Am Heart Assoc 10, e020085 (2021).

23. Seymour, C.W., et al. Derivation, Validation, and Potential Treatment Implications of Novel Clinical Phenotypes for Sepsis. Jama 321, 2003–2017 (2019).

24. Jeong, H., et al. Event-based contrastive learning for medical time series. arXiv preprint arXiv:2312.10308 (2023).

25. Weis, F., et al. Association between vasopressor dependence and early outcome in patients after cardiac surgery. Anaesthesia 61, 938–942 (2006).

26. Shahin, J., DeVarennes, B., Tse, C.W., Amarica, D.A. & Dial, S. The relationship between inotrope exposure, six-hour postoperative physiological variables, hospital mortality and renal dysfunction in patients undergoing cardiac surgery. Crit Care 15, R162 (2011).

27. Bellos, I., Iliopoulos, D.C. & Perrea, D.N. Association of postoperative fluid overload with adverse outcomes after congenital heart surgery: a systematic review and dose-response meta-analysis. Pediatr Nephrol 35, 1109–1119 (2020).

28. Delpachitra, M.R., Namachivayam, S.P., Millar, J., Delzoppo, C. & Butt, W.W. A Case-Control Analysis of Postoperative Fluid Balance and Mortality After Pediatric Cardiac Surgery. Pediatr Crit Care Med 18, 614–622 (2017).

29. Johnson, A.E.W., et al. MIMIC-IV, a freely accessible electronic health record dataset. Sci Data 10, 1 (2023).

30. Rodemund, N., Wernly, B., Jung, C., Cozowicz, C. & Koköfer, A. The Salzburg Intensive Care database (SICdb): an openly available critical care dataset. Intensive Care Med 49, 700–702 (2023).

31. Goradia, S., Sardaneh, A.A., Narayan, S.W., Penm, J. & Patanwala, A.E. Vasopressor dose equivalence: A scoping review and suggested formula. J Crit Care 61, 233–240 (2021).

32. Mathew, R., et al. Milrinone as Compared with Dobutamine in the Treatment of Cardiogenic Shock. N Engl J Med 385, 516–525 (2021).

33. Desman, J.M., et al. A distributional reinforcement learning model for optimal glucose control after cardiac surgery. NPJ Digit Med 8, 313 (2025).

34. Reimers, N. & Gurevych, I. Sentence-bert: Sentence embeddings using siamese bert-networks. arXiv preprint arXiv:1908.10084 (2019).

35. Paszke, A. Pytorch: An imperative style, high-performance deep learning library. arXiv preprint arXiv:1912.01703 (2019).

36. Ng, A., Jordan, M. & Weiss, Y. On spectral clustering: Analysis and an algorithm. Advances in neural information processing systems 14(2001).

37. Rousseeuw, P.J. Silhouettes: a graphical aid to the interpretation and validation of cluster analysis. Journal of computational and applied mathematics 20, 53–65 (1987).

38. Caliński, T. & Harabasz, J. A dendrite method for cluster analysis. Communications in Statistics-theory and Methods 3, 1–27 (1974).

39. Hotelling, H. Analysis of a complex of statistical variables into principal components. Journal of educational psychology 24, 417 (1933).

40. Van der Maaten, L. & Hinton, G. Visualizing data using t-SNE. Journal of machine learning research 9(2008).

41. Firth, D. Bias reduction of maximum likelihood estimates. Biometrika 80, 27–38 (1993).

42. Heinze, G. & Schemper, M. A solution to the problem of separation in logistic regression. Statistics in medicine 21, 2409–2419 (2002).

