## Supplement for "Contrastive Transformer-Driven Discovery of Temporal Hemodynamic Subphenotypes in Cardiac Surgery Patients"

### Table of Contents

Figures....................................................................................................................2

Tables.....................................................................................................................4

### Figures

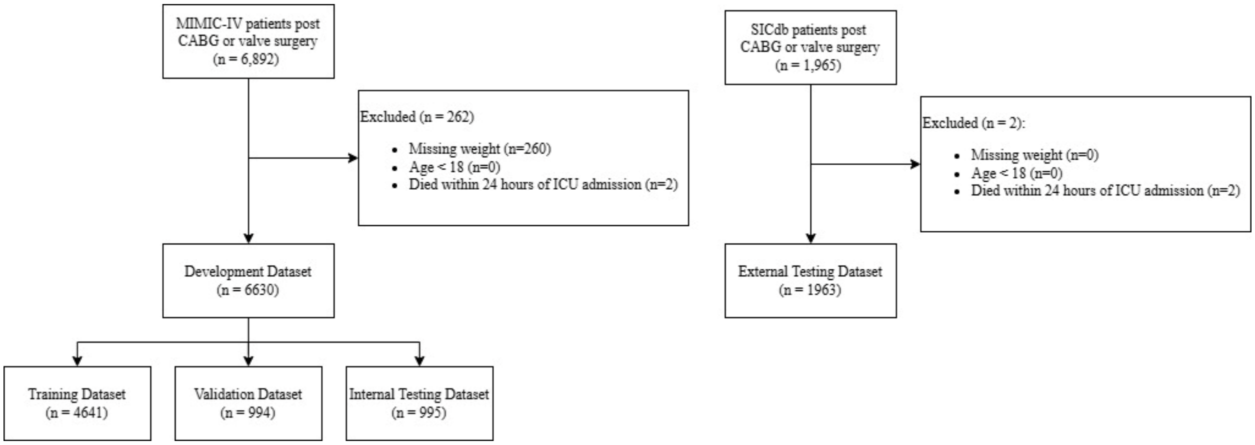


**Supplementary Figure 1**. CONSORT diagram of patient inclusion criteria.

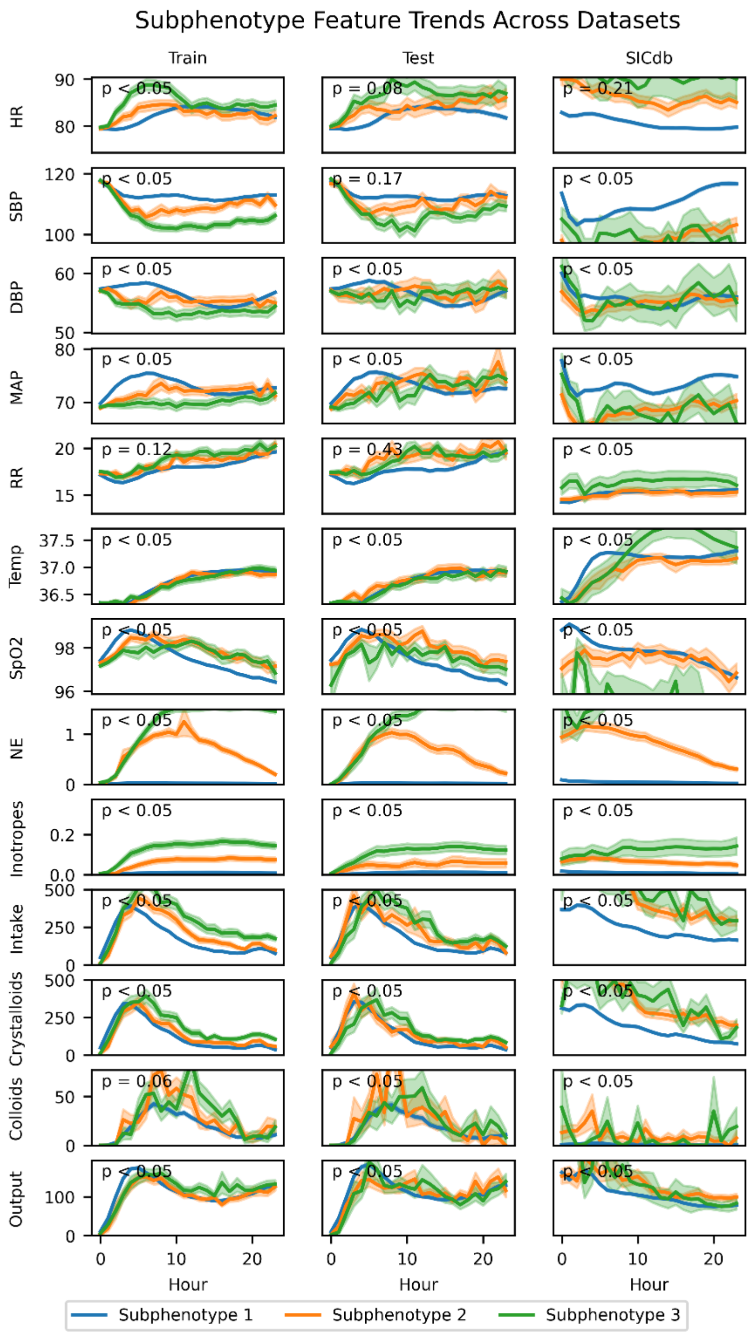


**Supplementary Figure 2**. Mean feature value ± standard error of the mean at each hour within each DTW-derived cluster. Linear mixed-effects modeling used to test whether the time-series trend for each clinical feature differed significantly across patient subphenotypes. NE = norepinephrine equivalent dose; Inotropes = milrinone equivalent dose

### Tables

**Supplementary Table 1. Clinical characteristics of patients assigned to each subphenotype**. Subphenotypes derived from the DTW-derived embeddings. S1 = subphenotype 1, S2 = subphenotype 2, S3 = subphenotype 3

| | **Training** | | | | **Testing** | | | | **External Validation** | | | |
|---|---|---|---|---|---|---|---|---|---|---|---|---|
| **Variable** | **S1** | **S2** | **S3** | **p-value** | **S1** | **S2** | **S3** | **p-value** | **S1** | **S2** | **S3** | **p-value** |
| Count (n) | 4390 | 123 | 128 | | 1889 | 48 | 52 | | 1883 | 67 | 13 | |
| Gender (% Male) | 72% | 76% | 74% | p = 0.53 | 72% | 73% | 75% | p = 0.89 | 73% | 66% | 77% | p = 0.35 |
| Admission Age (years) | 67.60 (11.37) | 69.02 (11.44) | 68.48 (11.77) | p = 0.28 | 67.69 (11.03) | 68.27 (12.71) | 66.71 (11.47) | p = 0.77 | 66.92 (11.08) | 69.18 (8.69) | 65.00 (18.26) | p = 0.21 |
| Mean Inotrope Rate (mcg/kg/min) | 0.01 (0.05) | 0.06 (0.11) | 0.13 (0.14) | **p < 0.01** | 0.01 (0.05) | 0.03 (0.08) | 0.09 (0.13) | **p < 0.01** | 0.01 (0.03) | 0.06 (0.08) | 0.12 (0.12) | **p < 0.01** |
| Mean NE Rate (mcg/kg/min) | 0.02 (0.03) | 0.66 (0.36) | 1.20 (0.46) | **p < 0.01** | 0.02 (0.03) | 0.51 (0.29) | 1.20 (0.37) | **p < 0.01** | 0.04 (0.07) | 0.81 (0.48) | 1.94 (0.55) | **p < 0.01** |
| Net Fluid Balance (mL) | 1514.62 (1809.07) | 2632.50 (1660.07) | 4071.26 (2779.79) | **p < 0.01** | 986.20 (1495.80) | 1519.44 (2247.79) | 2237.75 (1975.66) | **p < 0.01** | 3382.39 (2479.59) | 8826.20 (3499.03) | 8647.91 (5139.33) | **p < 0.01** |
| Total Intake (mL) | 4193.29 (1796.18) | 5117.24 (1853.71) | 6890.95 (3335.13) | **p < 0.01** | 2826.88 (2400.55) | 3351.73 (2943.46) | 4012.54 (3546.71) | **p = 0.02** | 5960.64 (2646.17) | 11976.91 (4322.26) | 12380.40 (5914.20) | **p < 0.01** |
| Total Crystalloid (mL) | 3137.73 (1346.70) | 3395.33 (1365.96) | 4489.71 (2103.97) | **p < 0.01** | 3203.79 (1374.11) | 3580.11 (1865.17) | 4043.65 (1525.66) | **p < 0.01** | 4398.63 (2380.35) | 9531.01 (3696.33) | 8936.52 (4283.94) | **p < 0.01** |
| Total Colloid (mL) | 447.88 (486.08) | 659.31 (562.45) | 708.98 (480.15) | **p < 0.01** | 446.51 (495.02) | 682.81 (578.60) | 555.88 (497.08) | p = 0.21 | 20.27 (108.19) | 240.82 (362.12) | 217.31 (359.05) | p = 0.07 |

**Supplementary Table 2. Univariable analysis of outcome characteristics of patients assigned to each subphenotype.** Subphenotypes derived from the transformer-derived embeddings. S1 = subphenotype 1, S2 = subphenotype 2, S3 = subphenotype 3.

| | **Training** | | | | **Testing** | | | | **External Validation** | | | |
|---|---|---|---|---|---|---|---|---|---|---|---|---|
| **Variable** | **S1** | **S2** | **S3** | **p-value** | **S1** | **S2** | **S3** | **p-value** | **S1** | **S2** | **S3** | **p-value** |
| **Count (n)** | 4390 | 123 | 128 | | 1889 | 48 | 52 | | 1883 | 67 | 13 | |
| **ICU LOS (days)** | 2.63 (3.41) | 5.08 (4.92) | 10.23 (11.51) | **p < 0.01** | 2.66 (3.80) | 5.29 (5.67) | 9.99 (9.12) | **p < 0.01** | 5.38 (7.04) | 13.11 (11.44) | 20.63 (20.72) | **p < 0.01** |
| **Hospital LOS (Days)** | 8.53 (5.41) | 14.26 (10.22) | 18.91 (14.01) | **p < 0.01** | 8.55 (5.74) | 12.11 (6.77) | 19.47 (12.04) | **p = 0.02** | 12.89 (9.91) | 23.37 (17.90) | 27.77 (23.92) | **p = 0.04** |
| **In-Hospital Mortality (% Died)** | 1% | 5% | 18% | **p < 0.01** | 0% | 0% | 12% | **p < 0.01** | 3% | 18% | 38% | **p < 0.01** |

**Supplemental Table 3. ICD-9-PCS and ICD-10-PCS codes**. Table of codes used to identify CABG, valve repair, and valve replacement procedures.

| Category | Code | Translation |
|---|---|---|
| CABG | 361X | Aortocoronary Bypass for Heart Revascularization |
| | 2100X | Coronary Artery, One Artery |
| | 2110X | Coronary Artery, Two Arteries |
| | 2120X | Coronary Artery, Three Arteries |
| | 2130X | Coronary Artery, Four or More Arteries |
| Valve Repair/Replacement | 352X | Open and other replacement of unspecified heart valve |
| | 3511 | Open heart valvuloplasty of aortic valve without replacement |
| | 3512 | Open heart valvuloplasty of mitral valve without replacement |
| | 3513 | Open heart valvuloplasty of pulmonary valve without replacement |
| | 3514 | Open heart valvuloplasty of tricuspid valve without replacement |
| | 02RF0X | Replacement, Aortic Valve |
| | 02RG0X | Replacement, Tricuspid Valve |
| | 02RH0X | Replacement, Pulmonary Valve |
| | 02RJ0X | Replacement, Tricuspid Valve |
| | 02QF0X | Repair, Aortic Valve |
| | 02QG0X | Repair, Mitral Valve |
| | 02QH0X | Repair, Pulmonary Valve |
| | 02QJ0X | Repair, Tricuspid Valve |

**Supplementary Table 4.** All model hyperparameters.

| **Hyperparameter** | **Value** |
|---|---|
| Transformer feedforward dimension | 512 |
| # Transformer Layers | 6 |
| # Transformer Heads | 4 |
| d_model | 64 |
| Latent dimension size | 64 |
| Latent dimension hidden layer size | 256 |
| Projection head hidden layer size | 256 |
| Temperature | 0.5 |
